# Rapid Neurotoxicity of Transfluthrin Volatiles and Spatial Repellency of Alternate Deterrents Against Host Seeking Black Flies, *Simulium Vittatum* (Diptera: Simuliidae)

**DOI:** 10.64898/2026.10.22.26366219

**Authors:** Bridget O. Aito-Bobadoye, Sklyer M. Kerr, Elmer W. Gray, Conor G. Fair, Darold P. Batzer

## Abstract

Spatial repellants are novel tools that aim to disrupt host seeking behavior in hematophagous insects. Outdoor spatial repellents succeed only when the aerodynamic channel is predictable enough to maintain a behavioral repellent field. Though standardized bioassays are essential for characterizing deterrent mediated behavioral responses, substantial knowledge gaps remain about the practical use of high throughput assays to evaluate repellency in outdoor biting species such as *Simulium* species. This is particularly relevant within the context of spatial repellent formulations for outdoor use. This study evaluated the comparative spatial repellency, knockdown and lethality thresholds of transfluthrin, permethrin, DEET (N, N diethyl meta toluamide), citronella oil and neem oil against host-seeking adult black flies (Diptera: Simuliidae). Concentration response data obtained (T=15, 30mins,1hr and 24hrs) across different concentration scales (0.001ug-10mg/cm^2^) were analyzed using a nonlinear regression to four parameter logistic equations to calculate half repellency (EC50), half knockdown (KC50) and half lethality (L50) concentration thresholds respectively. Results revealed that black flies exhibited significant repellency, knock down and lethality rates towards transfluthrin in horizontal orientation compared to DEET, permethrin, citronella and neem oil (χ² = 48.99, *df* = 32, P = 0.028). Driven by rapid vapor phase, transfluthrin demonstrated unparalleled repellency at a concentration of 0.1ug/cm^2^ in horizontal orientation (EC_50_ values = 0.0201 μg/cm²) and vertical orientation (EC_50_ values = 0.030 μg/cm²) compared to DEET (1h EC50 = 39.72 μg/cm²). 1h KC_50_ values revealed rapid knockdown effects (0.0259 μg/cm²) compared to DEET (38.24 μg/cm²) evidenced by rapid progression of neurotoxic hyper excitation in black flies in both orientations. Permethrin showed least repellency nor knockdown rates even at the highest concentrations (10mg/cm^2^). Compound concentration significantly impacted chemical diffusion dynamics (χ² = 75.85, *df* = 36, P = 0.000116), revealing that total compound loss through time depends heavily on chemical type. In conclusion, results from this study validates dose dependent spatial repellency of transfluthrin against the outdoor biting insect vector, *Simulium vittatum* in varied orientation placements using a high throughput repellency assay. This underscores the potential of spatial repellent compounds as a promising novel tool against both vectors of onchocerciasis, *Simulium damnosum s.l.* and vesicular stomatitis virus, *Simulium vittatum*.

## Introduction

Black flies belonging to the family Simuliidae (Diptera) represent a globally pervasive cohort of hematophagous insects possessing immense public health and veterinary significance (Adler et al., 2004; Adler et al., 2010; Adler and Crosskey 2019; Rivera-Martinez et al., 2025). *Simulium vittatum* serves as a competent vector of vesicular stomatitis virus, while *Simulium damnosum complex s.l and s.s* serves as both primary vectors responsible for the transmission of *Onchocerca volvulus*, the filarial nematode parasite causing human onchocerciasis, commonly known as river blindness (Boatin and Richards 2006). Beyond filarial transmission, the persistent, aggressive biting behavior of female *simuliids* inflicts severe systemic trauma on avian and mammalian hosts, culminating in a clinical state known as “black fly toxicosis” or “simuliotoxicosis” (Smith et al., 2009). Driven by potent, immunogenic salivary anticoagulants injected during blood-feeding bouts, severe allergic reactions can induce anaphylactic shock, and systemic toxemia in both livestock and wildlife (Shelley and Coscaron 2001). Conventional vector control strategies for *Simuliids* have heavily prioritized the chemical or biological larviciding of fast-flowing river systems where black fly larvae breed. However, this provides limited protection against hosts seeking adults in peri-urban outdoor spaces where animal or human-black fly contact frequently occurs such as camping sites or golf courses. As a result, there is growing interest in spatial repellents, also referred to as spatial emanators, which could drastically reduce host seeking and human landings in open or semi-open environments (Achee et al., 2012; Soderlund, 2012; White & Moore, 2015).

An insect repellent (also commonly called a “ deterrent”) is a substance applied to the skin, clothing, or other surfaces to discourage biting insects (arthropods in general) from landing or climbing onto that surface (White & Moore, 2015). Insect repellents are known to prevent insect-human contact and ultimately control the outbreak of insect-borne (and other arthropod-borne) diseases such as malaria, lyme disease, dengue fever, river blindness, and West Nile fever amongst several others. Depending on their mode of application, repellents are broadly categorized as spatial or topical (Achee et al., 2012). Spatial repellents (emanators) can complement existing vector control tools as they have increasingly been touted as a promising line of defense against biting arthropods (Achee et al., 2012; White & Moore, 2015). These spatial repellents function by releasing their volatile compounds into the immediate environment at pre-determined intervals, thereby creating a protective zone that can deter biting insects before contact occurs. In contrast, topical repellents are applied directly to exposed skin or clothing to provide personal protection against bites from blood-feeding arthropods (White & Moore, 2015). This interruption of host-seeking and human landing is the endpoint of all repellent products (spatial or topical), as commonly used compounds all aim at preventing human contact.

For topical repellents, the U.S. Environmental Protection Agency up till date has approved seven active ingredients, namely DEET, picaridin, IR3535, 2-undecanone, oil of citronella, oil of lemon eucalyptus, and catnip oil (EPA, 2018). Notwithstanding, several other plant-derived compounds including geraniol and linalool have shown significant promise (Bandi and Naravula 2026). These work by releasing volatile active ingredients that can interfere with host-seeking behavior, decrease landings, or form protective zones around users (human or animals). Interestingly, among these compounds, DEET, which is a contact repellent, continues to be regarded as the gold standard topical repellent because of its proven effectiveness against a broad range of mosquito species and other disease-causing arthropods (Leal, 2014). DEET belongs to the dialkylamide chemical class (specifically as an N, N dialkylamide or aromatic carbonamide) and it functions primarily at close range by interacting with an insect’s odorant receptors to block or mask host cues. However, increasing evidence now suggests that prolonged exposure to DEET may contribute to the development of behavioral resistance in mosquito populations (Pellegrino et al., 2011; Stanczyk et al., 2013; Sugiharto et al., 2016). Particularly as knockdown resistance (*kdr*) mutations are increasingly becoming widespread in wild mosquito populations (Wagman et al., 2015; Agramonte et al., 2017).

Numerous studies continue to characterize the neurotoxic effects of spatial repellent chemistry on the interruption in several disease vectors, especially mosquito species such as *Anopheles, Aedes* and *Culex* (Syed & Leal, 2008; Ogoma et al., 2012; Soderlund, 2012) under indoor settings. These studies have repeatedly reiterated the promising potential of spatial repellents in preventing the spread of mosquito-borne diseases. Therefore, the 2025 WHO guidelines for vector control now formally recognizes spatial repellents (emanators) as a recommended intervention class, noting that “effective protection of human populations with spatial repellents depends on continuous high coverage in the targeted area over an extended duration of time (WHO 2025). Nonetheless, the consumer market continues to offer a wide range of spatial repellent products such as ultrasonic devices, burning coils and diffusers, that differ greatly in their active ingredients, deployment type and delivery methods (WHO, 2015).

Over the past decade, a wide range of synthetic and natural compounds have been identified, characterized and developed as spatial repellents (Ogoma et al., 2012; Revay et al., 2013; White & Moore, 2015). Therefore, they have increasingly been incorporated into commercial repellent products such as mosquito coils, passive emanators, and dispensers (Ogoma et al., 2012; Revay et al., 2013; White & Moore, 2015). Common spatial repellent compounds include volatile synthetic pyrethroids such as allethrin, metofluthrin, and transfluthrin which have all demonstrated strong deterrent potential in both laboratory and semi-field tests because of their quick action and ability to disperse effectively in the air (Ogoma et al., 2012). Allethrin, transfluthrin and metofluthrin are type 1 synthetic pyrethroids, which are widely used mosquito adulticides and spatial repellents ( Xu et al., 2014). All three compounds, follow a core pyrethroid neurotoxic mechanism by binding to voltage gated sodium channels (VGSCs) of insect olfactory receptors, causing persistent sodium influx, repetitive firing, extreme hyperexcitation and finally knockdown ultimately leading to mortality (Syed & Leal, 2008; Xu et al., 2014). Common visible symptoms of neurotoxic hyper excitation in insects are full body tremors, spasms, loss of coordination, seizures, limb loss and eventual paralysis. Given the plethora of information on the wide use of spatial repellents for indoor biting insects especially mosquito species such as *Aedes aegypti* and *Culex pipiens* (Syed & Leal, 2008; Ogoma et al., 2012; Soderlund, 2012; Xu et al., 2014), there is a lack of evidence of similar compounds, particularly synthetic volatile pyrethroids, as effective repellents against aggressive outdoor biting insects such as Simuliids. However, because specific neuro-receptors for *Simulium* have not been fully mapped till date and remain poorly understood, it’s uncertain which odorant receptors in black flies are responsible for detection and avoidance.

This has spurned the need to validate reliable screening methods which can rapidly evaluate candidate compounds without the limitations associated with conventional behavioral bioassays. Existing methods, including olfactometers and choice-based assays, often require relatively large quantities of test compounds (milligram levels), involve complex experimental procedures, and can be difficult to decontaminate for repeated use (Rutledge et al., 2015). To address this methodological gap, this study validated a high throughput assay to characterize the comparative spatial repellency, knockdown and lethality dynamics of transfluthrin, permethrin, DEET, neem oil, and citronella oil against host seeking black flies, *Simulium vittatum*.

This method, which is a simple, rapid, and high-throughput bioassay method has already been used to evaluate spatial repellency and vapor toxicity (Jiang et al., 2019) of selected repellent compounds against the Orlando strain of *Aedes aegypti* (L.) (Diptera: Culicidae) under laboratory conditions. The screening apparatus has been designed to be inexpensive, compact, easy to decontaminate between replicates, and capable of producing behavioral responses using only microgram quantities of test compounds. We therefore hypothesized that the spatial synthetic pyrethroid (transfluthrin) would elicit distinct behavioral repellency, knockdown and lethality profiles (EC_50,_ KC_50_ and LC_50 values_) irrespective of orientation when compared with contact irritants, plant-based botanicals and topical repellents. The performance of this assay was assessed using representative compounds with different modes of repellency action, including the contact irritant permethrin, contact repellent DEET, plant based botanical neem and citronella oil, and the volatile spatial repellent transfluthrin. This is particularly relevant for black flies specifically, where river corridor airflow creates a unique diffusion channel that differs from indoor mosquito environments.

## Materials and methods

### Chemicals

DEET (98%) was purchased from Thermo Fisher Scientific (Waltham, MA, U.S.A), citronella oil (98% citronellol) was purchased from 1st Scientific, transfluthrin (99.9%) was purchased from Santa Cruz Biotechnology, permethrin (95%, mixed isomers) was purchased from AstaTech Incorporated, while Neem oil (99%) was sourced locally (**Fig.1**). Acetone (99.6%) for use as a solvent was also purchased from Thermo Fisher Scientific (Waltham, MA, U.S.A).

**Figure 1.**
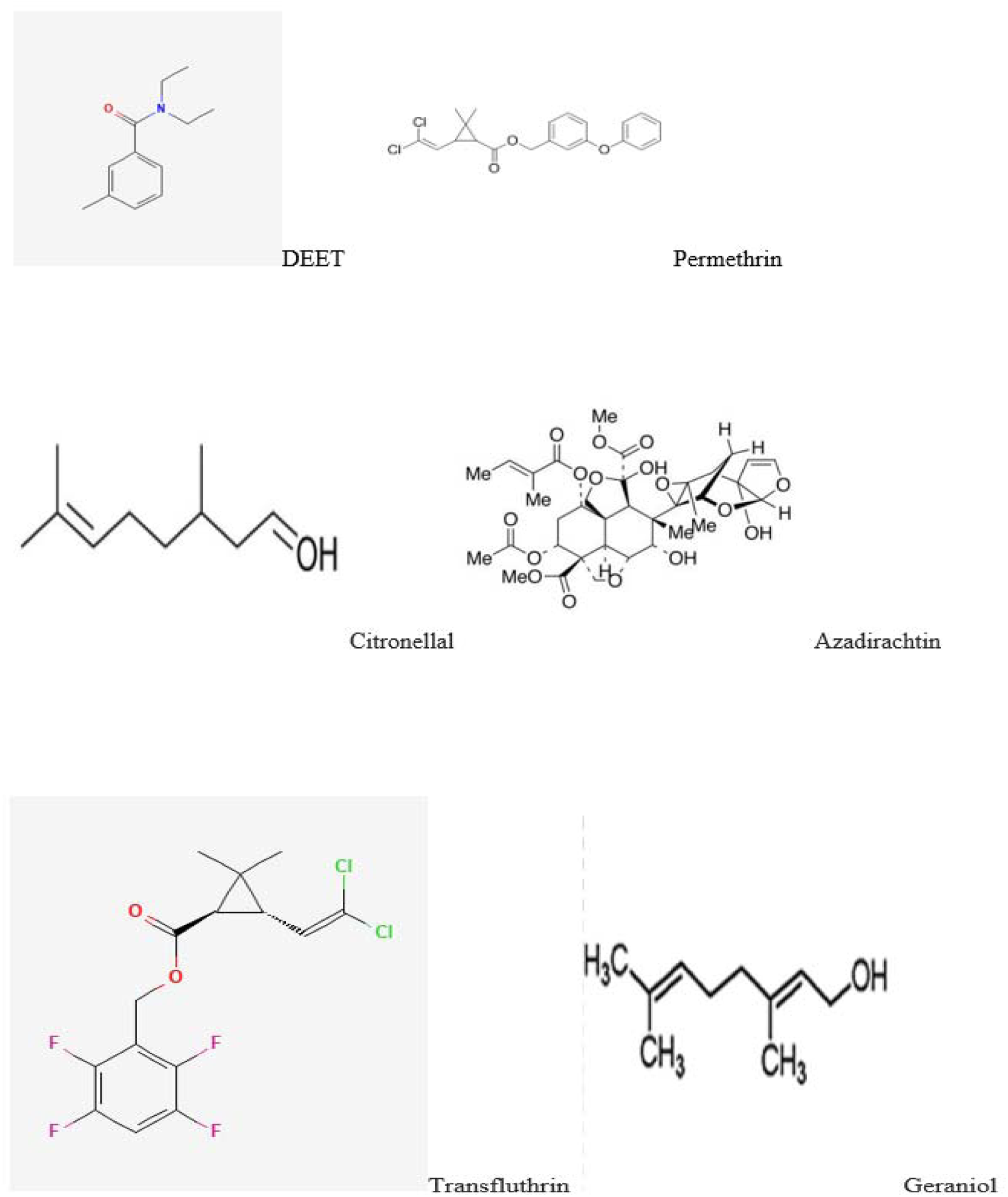
Chemical structures of the compounds tested. DEET, N, N-diethyl-meta-toluamide, citronella oil (citronellal and geraniol), Neem oil (Azadirachtin), permethrin, transfluthrin.

### Insects

Female black flies were obtained from the colony maintained at the black fly research and resource center (Athens, Georgia U.S.A.) and used for bioassays.

### Repellency assay

Adult *Simulium vittatum* flies that were at least 21 days old were selected and cooled at 20°C for 5 minutes using a slightly modified cold anesthesia technique (Chen et al., 2014). Subsequently, 20 females were transferred to each glass tube (length 12.5 cm, outer diameter 2.5 cm) (Tri- Kinetics Inc., Waltham, MA, U.S.A.). Within this time frame, the flies equally distributed throughout the glass tube without any obvious positional bias, after showing occasional bouts of walking, prior to recovery from cold induced anesthesia (**Fig. 2A**). Each glass tube was enclosed on each end by cloth netting held in place with blue end caps removed from 50-mL polypropylene centrifuge tubes (Falcon™; Corning Inc., Corning, NY, U.S.A.) The tubes were then placed on a white Styrofoam board with a black line drawn in the middle and were secured against rolling off by entomological pins attached to the board (**Fig. 2B-C**). Round filter papers were cut to size (diameter 2.5 cm) (Sigma-Aldrich Chemical Co.) and treated with 50 μL solution of compound at different concentrations dissolved already in acetone but were given additional 10 min for complete acetone evaporation (**Fig 2D**). The end caps bearing treated with filter paper of different concentrations were then affixed to the glass tubes, by replacing the blue caps with the netting held tightly in place with rubber bands. All end caps containing filter papers were positioned approximately 0.5 cm from the glass tubes to prevent black fly contact with the treatment filter papers. Control experiments were set up in a similar pattern with filter papers on both sides treated with 50 μL of acetone, or a DEET treatment at 100 μg/cm² serving as negative and positive controls, respectively. (**Figure 2E**). Placement of each tube on the Styrofoam board containing the treated filter were randomized using a random number generator (Barker and Kelsey 2015). Data were recorded at specific time points of 15 min, 30 min and 1 h for repellency and knockdown, as well as at 24 h for mortality. Each concentration was repeated on three different batches of black flies. After each replicate, the netting, filter paper and rubber bands were discarded, while the glass tubes were routinely decontaminated by washing in soapy water and rinsed with acetone and copious amounts of distilled water. Treatment caps were washed with soapy water and rinsed with methanol and distilled water. For the volatile spatial repellent (transfluthrin) which could stick to the glass tubing, all used treatment caps, netting and blue end caps were appropriately discarded after each experiment, and glass tubes were soaked overnight in methanol before carrying out routine decontamination to remove any remaining residue.

**Figure 2A:**
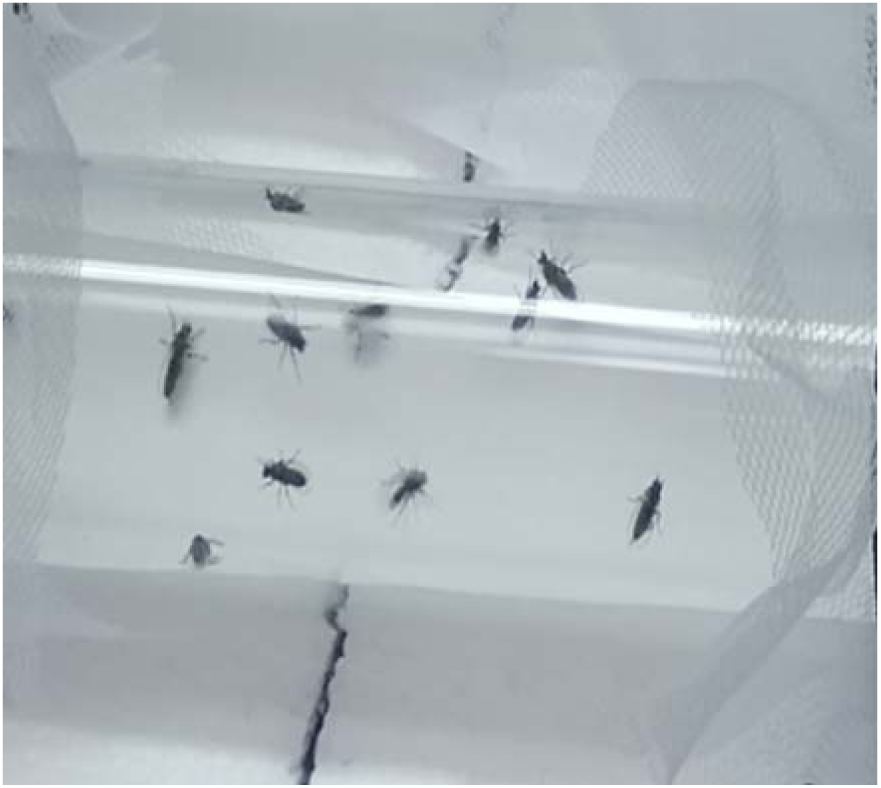
Flies recovery from cold induced anesthesia (cold narcosis) after 5 minutes.

**Figure 2.**
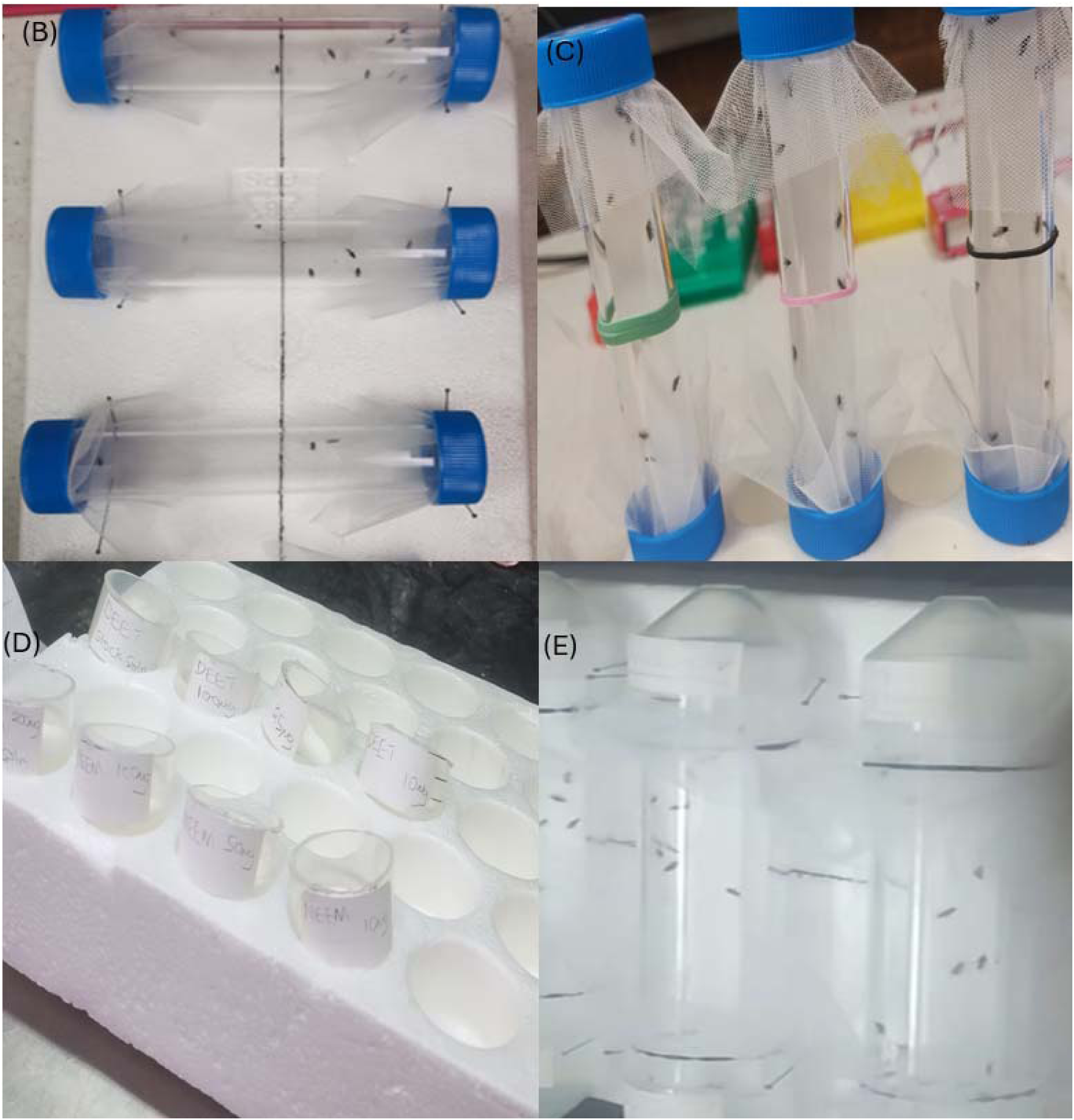
(B-E): Repellency test apparatus in horizontal (B) and vertical (C) orientation. Tube b and c show the holding caps, netting, filter paper caps and flies in both horizontal and vertical orientation prior to treatment. D shows the round treated filter papers (diameter 2.5 cm) (Sigma-Aldrich Chemical Co.) containing 50 μl solution of compound at different concentrations (0.001, 0.01, 0.1, 10, 25, 50 and 100ug/cm^2^). Tube E shows the control (positive and negative) experiments set up in a similar pattern with filter papers on both sides treated with 50 μL of acetone, or a DEET treatment at 100 μg/cm² serving as negative and positive controls, respectively.

To evaluate if orientation influences repellency outcomes in *Simulium vittatum*, the vertical placement was tested concurrently with the horizontal placement for comparative performance. Because black flies were unable to rest on the smooth vertical glass surface, a strip of filter paper (12.5 × 3 cm) (Chromatography paper, Fisher Scientific, Pittsburgh, PA, U.S.A.) was affixed inside the glass tube to provide a suitable resting substrate. Given that the inner circumference of the glass tubes measured approximately 6 cm, this dimension of filter paper was sufficient to cover roughly half of the inner surface area, thereby preserving the same proportional exposure used in the horizontal assay. The assembled glass tubes were held in a vertical orientation using Styrofoam boards originally designed to accommodate 50-mL centrifuge tubes, with all treated filter papers positioned at the bottom of the tube and the acetone control placed at the top. The positive control followed similar arrangement. Finally, a rubber band was affixed to the midline of each glass tube to enable quantification of the percentage of flies repelled by all treatment compounds following the same procedure described for the horizontal orientation assay.

### Chemical Evaporation Study

The evaporation rates of transfluthrin, DEET, permethrin, neem and citronella oil were evaluated at different time points to determine the diffused quantity and rates of each compound released from the filter papers. Circular filter papers were placed inside polypropylene caps and initially weighed using an analytical balance with a precision of 0.01 mg. 50 μL of the stock solution, as well as different concentrations diluted with acetone were applied to each filter paper. The filter papers were reweighed at predetermined time points of 15, 30, 60 minutes and 24 hours. The remaining amount of compound was calculated by subtracting the final measured weight from the initial weight. Each treatment concentration (0.001, 0.01, 0.1, 10, 25, 50, 100ug/cm^2^ and stock) were tested in three separate replicates for all chemicals (DEET, permethrin, transfluthrin, citronella and neem oil).

### Statistical analysis

Concentration-response analyses were conducted separately for repellency at 15 min (EC_50_), knockdown at 30 and 60 min (KC_50_), and mortality at 24 h (LC_50_). Responses were analyzed using quasibinomial logistic regression with chemical, tube orientation, log10-transformed concentration, and their interactions as predictors. EC_50_, KC_50_, and LC_50_ values and 95% confidence intervals were estimated from the fitted concentration-response relationships. Estimates occurring within this range were considered supported interpolations, whereas estimates above or below the tested range were identified as extrapolated and interpreted cautiously. Positive concentration-response relationships with limited statistical support were retained but identified as weak; 50% response concentrations were not estimated for non-increasing relationships. Numerical estimates were therefore interpreted together with their confidence intervals, fitted slopes, and estimate classifications. Controls were analyzed separately. Cumulative active-compound loss was analyzed using a linear mixed-effects model with chemical, concentration class, time, and their interactions as fixed effects and diffusion unit as a random effect. Post-15-min incremental loss was analyzed using a generalized least-squares repeated-measures model. Comparisons were based on estimated marginal means with multiplicity adjustment. Analyses were conducted in R version 4.5.1.

## Results

### Repellency in the horizontal tube orientation

Concentration-response relationships varied among chemicals, orientations, and assessment times **(Figs. 3A-3C**; **Tables 1-2)**. All tested compounds exhibited concentration-dependent repellency, each separately producing the expected sigmoidal concentration–response relationship with positive-slop extrapolated estimates **(Figure 3A)**. A concentration of 10μg/cm² and 100μg/cm² represented both the minimum and maximum thresholds required to elicit repellency, knock down and mortality in the flies. Transfluthrin generally produced responses at substantially lower concentrations than the other compounds, with several EC_50_, KC_50_, and LC_50_ estimates occurring within its tested concentration range. Transfluthrin induced repellency at 15- mins with EC_50_ values of 0.02018μg/cm², and at 1hr (0.02599ug/cm^2^), notably characterized with rapid muscle excitability and trembling in over 90% of flies assessed across all 3 treatment replicates. DEET produced repellency of 1hr EC values 38.24 μg/cm², which was not significantly different from, the 15-minute EC value of 39.72 μg/cm², reflecting DEET’s potency remaining stable over time, which further substantiates unchanged vapor concentration was maintained in this orientation **(Figure 3B)**. Citronella oil exhibited a 15-minute EC50 value of 167.3 μg/cm² and a 30-minute EC value of 182.7 μg/cm², indicating a weaker repellency strength compared to DEET and transfluthrin (Table 1). In contrast, neem oil and permethrin estimates were weakly supported or required extrapolation beyond the tested concentrations, and some concentration-response relationships were non-increasing and therefore did not yield estimable 50% response concentrations. Unlike transfluthrin, the contact irritant permethrin failed to demonstrate either significant repellency or knockdown responses in the flies following 1 hour of exposure, even at the highest concentrations of 10 mg/cm².

**Figure 3A:**
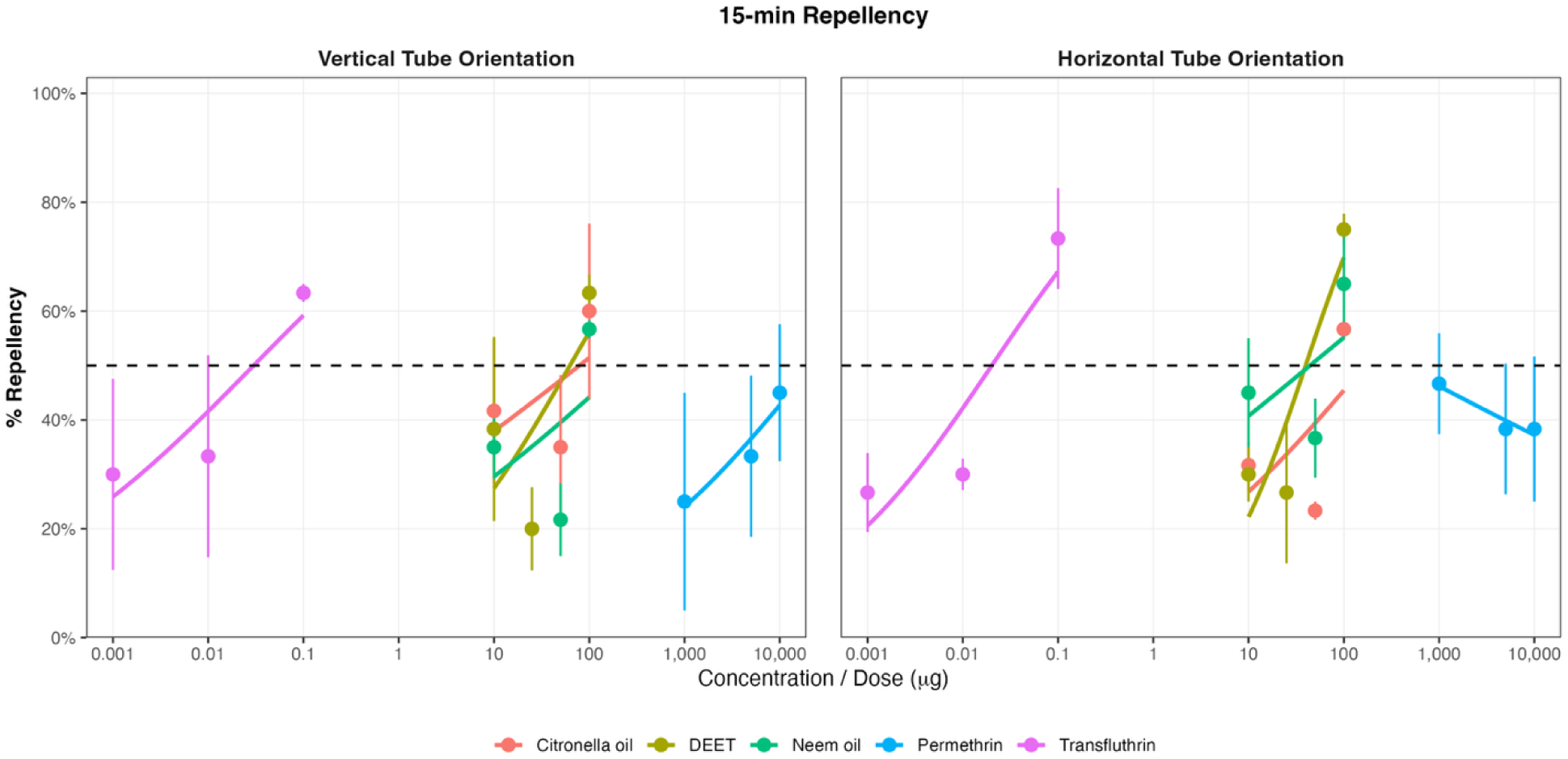
Concentration-response relationships for repellency at 15 min. Mean percentage repellency (±SE) of flies exposed to citronella oil, DEET, neem oil, permethrin, or transfluthrin in vertical and horizontal tube orientations. Lines represent fitted concentration-response relationships from quasibinomial regression models. The horizontal dashed line indicates 50% repellency, from which EC_50_ values were estimated. Concentrations are shown on a log_10_ scale.

**Figure 3B:**
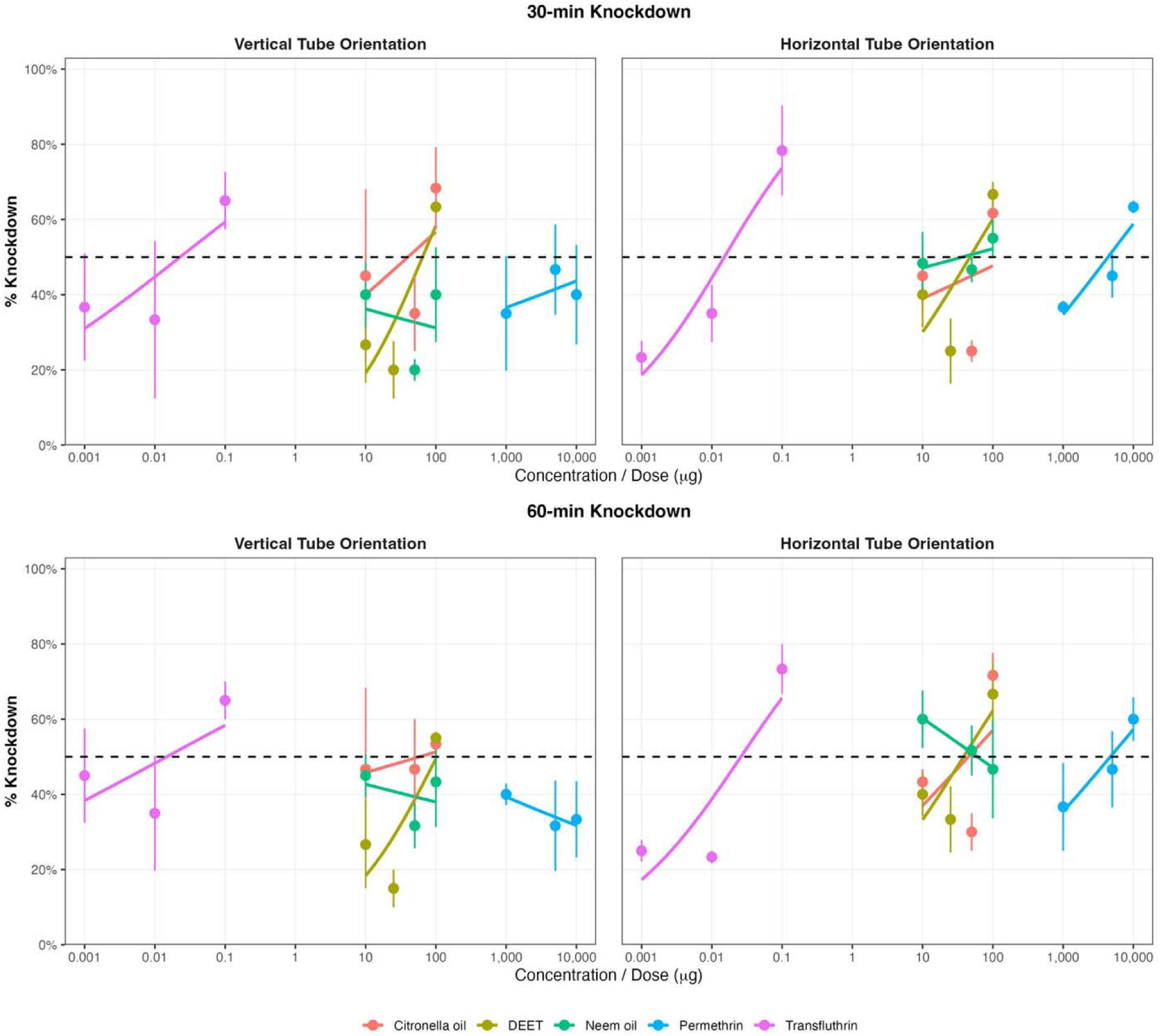
Concentration-response relationships for knockdown at 30 min and 60 min. Mean percentage knockdown (±SE) of flies exposed to citronella oil, DEET, neem oil, permethrin, or transfluthrin in vertical and horizontal tube orientations at 30 min (top) and 60 min (bottom). Lines represent fitted concentration-response relationships from quasibinomial regression models. The horizontal dashed line indicates 50% knockdown, from which KC_50_ values were estimated separately at 30- and 60-min. Concentrations are shown on a log_10_ scale.

**Figure 3C:**
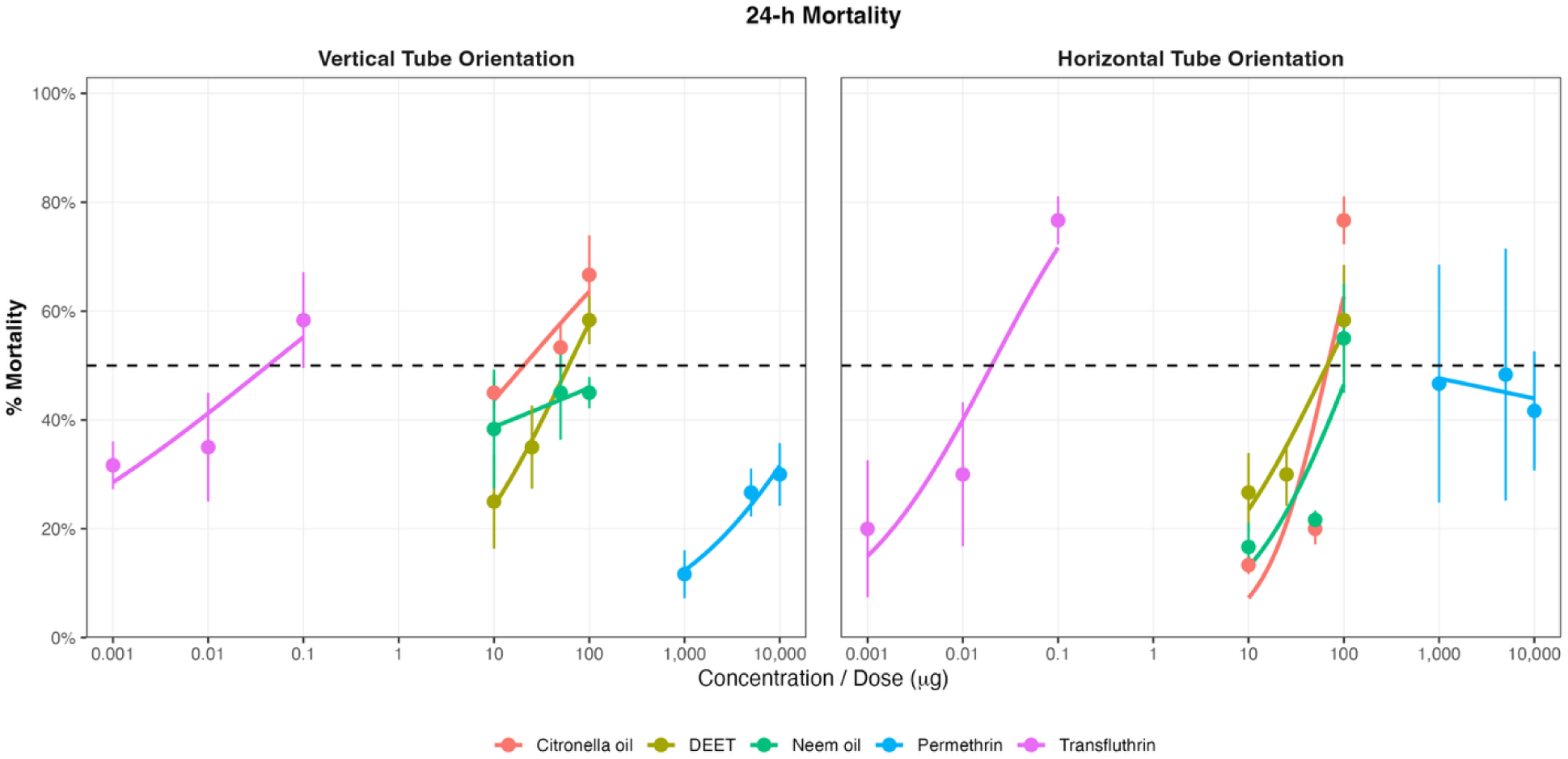
Concentration-response relationships for mortality at 24. **h.** Mean percentage mortality (±SE) of flies exposed to citronella oil, DEET, neem oil, permethrin, or transfluthrin in vertical and horizontal tube orientations. Lines represent fitted concentration-response relationships from quasibinomial regression models. The horizontal dashed line indicates 50% mortality, from which LC_50_ values were estimated. Concentrations are shown on a log_10_ scale.

**Table 1.** Estimated concentrations producing 50% repellency (EC□□) at 15 min for each chemical and tube orientation. EC□□ estimates and 95% confidence intervals were derived from quasibinomial concentration–response models using log**□□**-transformed concentration. Slope represents the fitted concentration-response slope, with P values testing whether the slope differed from zero. Supported interpolations occurred within the tested concentration range. Estimates outside the tested range are identified as extrapolated, whereas positive relationships with limited statistical support are identified as weak concentration-response slopes. EC□□ values were considered not estimable when the fitted concentration-response slope was non-increasing. Concentrations are expressed in µg.

| Orientation | Chemical | Estimate (95% CI) | Slope | P-value |
| --- | --- | --- | --- | --- |
| Horizontal | Citronella oil | 167.3 (7.982, 3507) | 0.8201 | 0.2691725 |
| Horizontal | DEET | 39.72 (19.49, 80.91) | 2.0941 | 0.0097713 |
| Horizontal | Neem oil | 44.15 (4.381, 445) | 0.5803 | 0.4030392 |
| Horizontal | Permethrin | Not estimable | -0.3692 | 0.5949242 |
| Horizontal | Transfluthrin | 0.02018 (0.004801, 0.08484) | 1.0349 | 0.0120849 |
| Vertical | Citronella oil | 78.9 (3.726, 1671) | 0.5473 | 0.4329337 |
| Vertical | DEET | 62.71 (15.52, 253.4) | 1.2186 | 0.1006613 |
| Vertical | Neem oil | 235.1 (2.332, 2.369e+04) | 0.6313 | 0.3855677 |
| Vertical | Permethrin | 2.19e+04 (772, 6.214e+05) | 0.8588 | 0.2595998 |
| Vertical | Transfluthrin | 0.03 (0.003422, 0.2629) | 0.7125 | 0.0628947 |

**Table 2.**
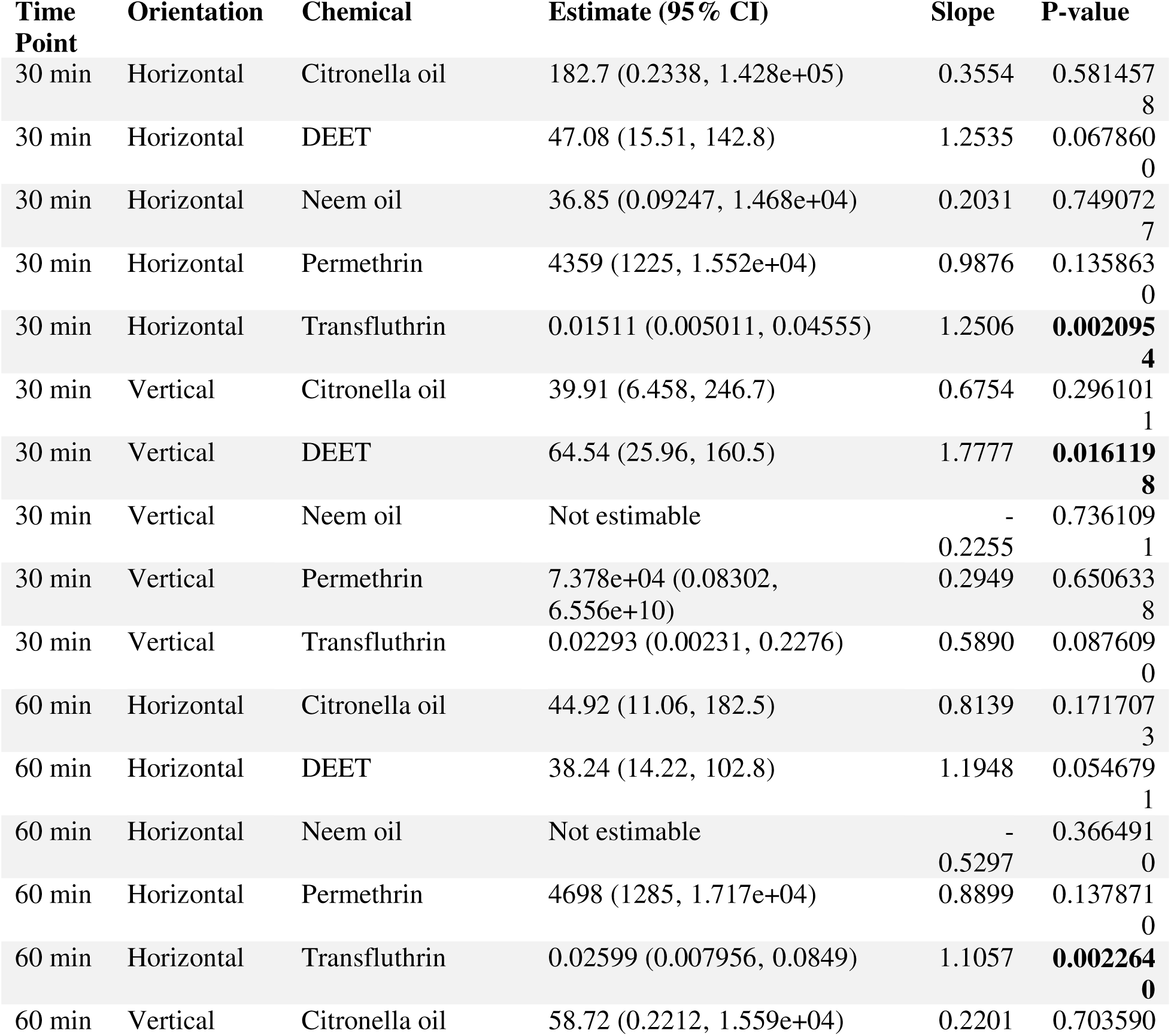
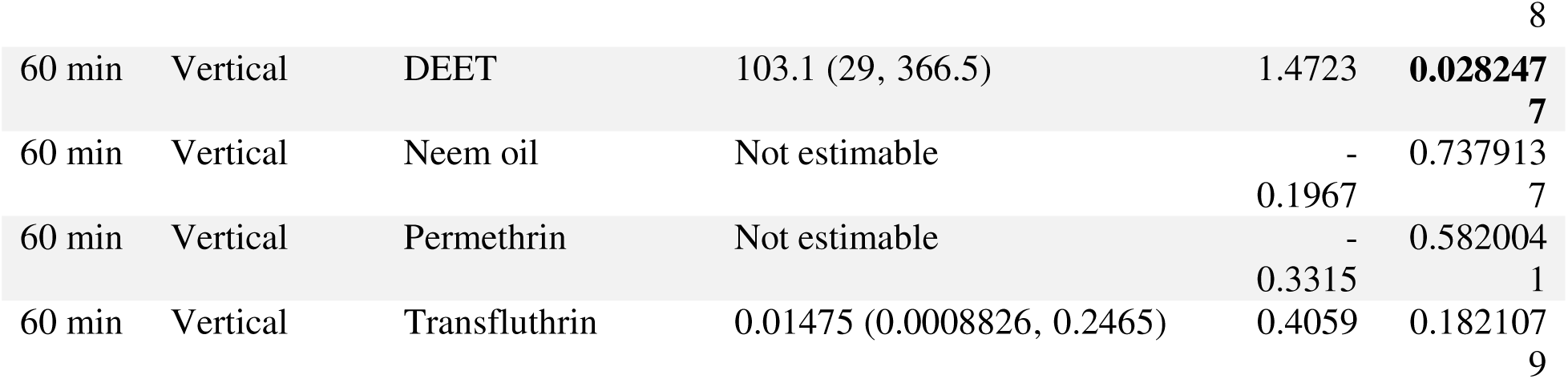
Estimated concentrations producing 50% knockdown (KC□□) at 30 and 60 min for each chemical and tube orientation. KC**□□** estimates and 95% confidence intervals were derived from quasibinomial concentration–response models using log□□-transformed concentration. Slope represents the fitted concentration-response slope, with P values testing whether the slope differed from zero. Supported interpolations occurred within the tested concentration range. Estimates outside the tested range are identified as extrapolated, whereas positive relationships with limited statistical support are identified as weak concentration-response slopes. KC□□ values were considered not estimable when the fitted concentration-response slope was non-increasing. Concentrations are expressed in µg.

**Table 3.** Estimated concentrations producing 50% mortality (LC□□) at 24 h for each chemical and tube orientation. LC□□ estimates and 95% confidence intervals were derived from quasibinomial concentration–response models using log□□-transformed concentration. Slope represents the fitted concentration-response slope, with P values testing whether the slope differed from zero. Supported interpolations occurred within the tested concentration range. Estimates outside the tested range are identified as extrapolated, whereas positive relationships with limited statistical support are identified as weak concentration-response slopes. LC□□ values were considered not estimable when the fitted concentration-response slope was non-increasing. Concentrations are expressed in µg.

| Chemical | Orientation | Estimate (95% CI) | Slope | P-value |
| --- | --- | --- | --- | --- |
| Citronella oil | Horizontal | 67.74 (44.19, 103.8) | 3.0591 | 0.0008009 |
| Citronella oil | Vertical | 20.59 (4.361, 97.23) | 0.8116 | 0.1560363 |
| DEET | Horizontal | 66.69 (24.45, 181.9) | 1.4304 | 0.0220662 |
| DEET | Vertical | 60.77 (23.66, 156.1) | 1.4563 | 0.0196115 |
| Neem oil | Horizontal | 120.2 (44.07, 327.8) | 1.7602 | 0.0173662 |
| Neem oil | Vertical | 360.8 (0.02986, 4.361e+06) | 0.2947 | 0.6036124 |
| Permethrin | Horizontal | Not estimable | -0.1490 | 0.7899066 |
| Permethrin | Vertical | 4.482e+04 (2171, 9.252e+05) | 1.1868 | 0.1178555 |
| Transfluthrin | Horizontal | 0.02012 (0.007832, 0.05168) | 1.3315 | 0.0003709 |
| Transfluthrin | Vertical | 0.04251 (0.003813, 0.4739) | 0.5634 | 0.0652396 |

### Repellency in the vertical tube orientation

To account for the diffusion behavior of transfluthrin, a compound which is denser than air, and requires time to permeate upward or disperse evenly throughout a three-dimensional space, we incorporated a strip of filter paper to the inner wall of all glass tubes, which was evaluated as the experimental arena **(Figure 2C)**. In comparison to the horizontal arrangement, the vertical set-up produced non-significant differences in concentration-dependent repellency response for transfluthrin **(Figure 3C**; **Table 1)** with 15-minute EC value = 0.030μg/cm², 1-hour ( 0.0148μg/cm² ) with no contrasting differences in observable repellency in the flies at both time points. Regardless, a rapid neurotoxic effect characterized by rapid muscle hyperexcitability and trembling was similarly observed in the flies exposed to treatment concentrations of 0.1μg/cm² from 15 mins. The remaining reference repellent compounds were similarly assessed, at 1-hr KC_50_, DEET (103.1μg/cm²) displayed EC values that were at least three-fold higher than those obtained in the horizontal configuration (P < 0.05), indicating weaker repellency activity in vertical orientations. In contrast, neem oil demonstrated lower lethality thresholds in the flies assessed under this orientation, with LC_50_ values of 360.8μg/cm². Consistent with the findings from the horizontal set-up, permethrin showed neither significant spatial repellency at 15mins (EC value = 21,903μg/cm²) or 24hr lethality activity (LC_50_ value = 44,821μg/cm²) even at concentrations of 10 mg/cm². It is noteworthy to mention that these non-estimable values fall outside the tested concentration range and should be interpreted cautiously. However, among all compounds evaluated, transfluthrin still demonstrated to be the most potent repellent in both orientations, exhibiting approximately over a 1,000-fold greater potency, knockdown and lethality than DEET at 1 hour of exposure.

### Vapor Toxicity

Knockdown after 1 hour of exposure and mortality after 24 hours were assessed across all experimental treatments and replicated 3 times. At concentrations approximating their respective EC_50_ values, neem oil, citronella oil, and permethrin did not induce knockdown within the first hour and produced only minimal mortality after 24 hours, irrespective of tube orientation. At 30 minutes, DEET showed very similar KC□□ values across both orientations: horizontal (47μg/cm^2^) and vertical (64μg/cm2), demonstrating DEETs stability over this length of time.

Unlike the other compounds, transfluthrin exhibited marked toxicity even at 60 minutes in both orientations (Table 2). Exposure to concentrations of 0.1 μg/cm² resulted in extensive body tremors within 15 minutes in both vertical and horizontal orientations. Transfluthrin achieved similar 50% knockdown thresholds (1-hr KC50 values = 0.0148 μg/cm²) in the vertical orientation compared to (1-hr KC50 values = 0.0259 μg/cm²) in horizontal orientation.

### Evaporation assay

The influence of concentration on diffusion rates was further investigated using stock compounds and serial dilutions of the test compounds. The test compounds were applied to all filter papers as solutions prepared in acetone. For most compounds tested, the estimated cumulative active-compound mass loss after 24 hours revealed that the lowest concentrations had the highest mass loss rates with citronella oil (10 μg/cm² = 0.500); DEET (10 μg/cm² = 0.567), neem oil (10 μg/cm²= 0.833); Permethrin (1mg/cm² =1.467); and transfluthrin (0.001μg/cm² = 0.233). This showed that cumulative mass loss depends largely on combinations of variables such as chemical concentration, and time. This trend was particularly evident for citronella oil, where the lowest concentration tested (10ug) produced significant mass loss rates at 24hr time point. Transfluthrin exhibited no significant evaporative loss after both 1hr and 24 hours of exposure at a concentration of 0.1μg/cm2. Similarly, acetone dilutions of DEET displayed only minimal evaporation, retaining 95 ± 1% of its initial weight after 24 hours at 100 μg/cm^2^. In contrast, permethrin demonstrated substantial evaporation, accounting for 73 ± 1% and 71 ± 1% of the initial weight at 1 hour and two-thirds of final weight lost at 24 hrs respectively (34 ± 2% and 26 ± 2%).

## Discussion

All compounds evaluated in this study were selected to represent different mechanisms of repellency ranging from spatial repellents, contact irritants or repellents, and botanical deterrents. The high through-put bioassay used in this study (Jiang et al., 2019) validates it’s use as a simplified screening system for repellent compounds against outdoor biting insects such as *Simulium vittatum*. Although conceptually like the airborne repellent assay developed for *Drosophila melanogaster* (Kwon et al., 2010), this assay proves to be a less technical prototype of the high-throughput assay as described by Grieco et al. (2005). The performance of this apparatus is consistent with findings reported by Jiang et al., 2019 as previous studies recorded approximately 50% repellency for DEET at concentrations of 48μg/cm², respectively, after 10 minutes of exposure (Grieco et al., 2005; Deletre et al., 2013). Similarly, our study produced biologically relevant estimates (1hr KC□□ values of 38.24μg/cm² for DEET) and (1hr KC□□ values of 0.025μg/cm² for transfluthrin) in horizontal tube orientations and (1hr KC□□ values of 103.1μg/cm² for DEET) and (1hr KC □□ values of 0.014μg/cm² for DEET) (Table 2). Consistent with findings from previous studies, permethrin failed to exhibit spatial repellency and this may point to increased insecticidal resistance to this pyrethroid, as no knockdown with only minimal mortality were observed, even at concentrations as high as 10 mg/cm², indicating that toxic effects of permethrin maybe largely contact-dependent at very high doses (Achee et al., 2009). Similar repellent behavioral responses to DEET have also been confirmed using other types of repellency assays, such as wind tunnel experiments, where EC□□ values ranged from 16 to 31 μg/cm² (Sharpington et al., 2000; Rutledge & Gupta, 2004; Tsikolia et al., 2013). The 1-hour EC□□ value of 38.24μg/cm² obtained with the present assay in the horizontal tube orientation validates that this high throughput assay can provide reliable behavioral responses comparable to established methods even for different targeted disease vectors such as outdoor host seeking *Simuliid* flies.

Differences in glass tube orientation significantly influenced the response of all repellent compounds evaluated in this study, although transfluthrin exhibited similar activity in both horizontal and vertical configurations. Permethrin, neem and citronella oil were less potent in the vertical assay, with EC□□ values approximately three-fold higher than those obtained in the horizontal tube orientation. Behavioral deterrence induced by DEET is primarily mediated through insect odorant receptors (Ors) (Syed & Leal, 2008; Xu et al., 2014), however, since the exact odorant receptor sequence for black flies (family Simuliidae) remains uncharacterized, it’s unknown if they use similar neuroreceptors such as is found in *Drosophila* (such as Ir40a) and mosquitoes’ species like *Aedes aegypti* (AaegOR54) (Wang et al., 2009; Pellegrino et al., 2011). In contrast, the universal mechanism of behavioral deterrence and spatial repellency by transfluthrin is primarily mediated through the activation of voltage-gated sodium channels (VGSC) rather than olfactory receptors as evidenced in DEET (Bohbot and Dickens, 2012; Obermayer, 2015; Dong, 2017). Therefore, it is expected that the same mechanisms that govern behavioral deterrence will hold true for black flies.

In contrast, the repellent activity of citronella oil which has been linked to citronellal, which is its major constituent of this oil strongly activates transient receptor potential A1 (TRPA1) channels in *Anopheles gambiae* (Wang et al., 2009; Kwon et al., 2010). In *Anopheles gambiae*, the malaria mosquito, transient receptor potential A1 (TRPA1) is a heat-activated ion channel expressed in distal antennal structures and plays a crucial role in peripheral thermoreception, helping adult females to detect and respond to temperature gradients during close-range host-seeking (Wang et al., 2009). Because finding a host via heat signatures is essential for malaria vectors, AgTRPA1 now commonly functions as a potential molecular target for new repellents designed to disrupt mosquito host-seeking behavior, however, this irritant receptor and its exact genetic pathways remain unmapped for black fly species. In the present study, concentration-dependent spatial repellency was demonstrated by all compounds except permethrin, whose relatively low vapor pressure limited both its vapor toxicity and its ability to function as an effective repellent at even high concentrations (Achee et al., 2009; Nentwig et al., 2017).

The responses observed in the horizontal and vertical assays indicate that compound diffusion dynamics works in synergy with concentration and volatility of the compound. Evidence for this was provided by separate trials which contained no filter papers (vertical orientations). The persistence of the highest concentration of transfluthrin residues on the glass tube surface caused rapid neurotoxicity ranging from whole body tremors and muscle spasms, suggesting that the behavioral and toxic effects of transfluthrin resulted from both vapor and contact exposure. Further evidence of compound distribution was obtained from the filter paper liner experiments, where in the vertical tube orientation, the filter paper restricted vapor movement and toxicity of transfluthrin. Black flies were observed to consistently avoid the lower portion of the tube with the highest concentration transfluthrin treated filter papers, supporting our notion that heavy molecular compounds at high concentration-maintained constant vapor toxicity over time with higher concentration gradients accumulating more near the bottom of the glass tube (treatment/ positive control). The similar knockdown and lethality thresholds produced by transfluthrin is largely due to its nature, being a heavier compound which is much denser than air. This suggests that a greater proportion of transfluthrin vapors diffused at a slower rate while adhering to the bottom portion of the glass surface while the paper liner provided the black flies with adhesive support, significantly contributing to similar diffusion rates.

In contrast, citronella oil evaporated rapidly, losing approximately 25% of its content after 1 h and about 70% after 24 h. This rapid loss is consistent with its relatively high volatility and vapor pressures of its principal constituents, citronellal (0.25 mmHg) and geraniol (0.03 mmHg) at 25 °C. This further explains its short window period of repellent effectiveness as reported previously by Trongtokit et al., 2005, which will necessitate timed re-applications if used as a contact repellent. Although neem oil has a vapor pressure (1.1 × 10□³ mmHg at 20 °C) slightly less comparable to that of citronella oil, its lower efficacy in this present assay is likely attributable to its low volatility because it is primarily composed of heavy fatty acids and complex triterpenoids, its reduced repellent activity could have possibly resulted from its low diffusion rates. Permethrin, by comparison, possesses an extremely low vapor pressure (5.2 × 10□□ mmHg at 25 °C) and therefore exhibited poor performance in this assay, which is consistent with its established role as a contact irritant rather than a spatial repellent (Achee et al., 2009).

## Conclusions

Our findings demonstrate that orientation and placements of repellent compounds exert a substantial influence on their diffusion kinetics and biological activity of volatile insecticidal compounds. Horizontal tube placement restricted air movement and promoted the pooling of dense transfluthrin vapors near the glass tube, thereby creating a concentrated repellent zone. In contrast, vertical orientation facilitated greater air circulation and dispersion, but transfluthrin dense vapors still encouraged pooling and vapor accumulation at the treatment sides. The orientation-driven difference produced a two-fold shift in transfluthrin KC□□ values, which underscores how vapor density and gravitational settling shape exposure pathways for highly volatile neurotoxic agents.

These results highlight the critical role of compound volatility, vapor density, and airflow dynamics in determining the efficacy of spatial repellents, especially synthetic pyrethroids for use in outdoor settings. Outdoor settings are known to create anisotropic diffusion (directional, uneven distribution patterns), therefore outdoor spatial repellents can succeed only when the aerodynamic channel is predictable enough to maintain a behavioral repellent field. Several diffusing materials can influence controlled release of repellent volatiles such as hessian fabrics (passive emanation), polymer matrices (controlled release), or wicks and pads (capillary diffusion).

Each substrate used as a dispensing medium can create a distinct diffusion geometry, ranging from circular, directional, pulsed, or layered. This may place emphasis factors such as placement orientation, combined with compound concentration, and structural materials (diffusing substrates) must be explicitly considered when designing and evaluating repellent compounds, especially for outdoor or semi-open environment use. This unravels new avenues for incorporating spatial repellent formulations such as transfluthrin or metofluthrin for outdoor control of host seeking black flies such as *Simulium vittatum* and *Simulium damnosum s.l and s.s* near riverbanks or semi-outdoor settings as novel tools are sought to suppress their populations.

## Acknowledgements

The *Simulium vittatum* used in this work were produced with the support of NIH Task Order C- 08, Contract No. HHSN2722017000351, Task Order No. 75N93020F00002 and obtained through BEI Resources, NIAID, NIH: *Simulium vittatum*, Adults NR-53893.

## Author contributions

BA, DB, EWG, and SMK conceptualized the experimental design. BA and SMK carried out experimental bioassays and data acquisition, CF carried out the analysis and interpretation of data for the work. BA drafted the manuscript. BA, DB, EWG, and SMK edited the manuscript.

## Data availability

Data is available upon request from the corresponding author.

## Notes

### Competing Interest Statement

The authors have declared no competing interest.

